# Knowledge, attitudes, practices and epidemiological profile of Muslim faithful receiving Covid-19 vaccines in Yaounde,Cameroon

**DOI:** 10.1101/2022.06.12.22276300

**Authors:** Adidja Amani, Efietngab Atembeh-Noura, Cheuyem Lekeumo Fabrice Zobel, Moussa Souaibou, Joseph Kamgno, Bissek Anne Cecile

## Abstract

In the response strategy against the Covid-19 pandemic, Cameroon has used to date, two types of vaccine namely: Sinopharm, Covishield/AstraZeneca. The objective of this study is to determine the KAP and the epidemiological profile of Muslim faithful receiving the Covid-19 vaccine at Mosque Biyem-Assi, Yaounde-Cameroon. Data on COVID-19, vaccination, and clinical characteristics were collected. A total of 58 participants took part in the first survey and equally received their first dose of the COVID-19 vaccine of their choice. The majority of the participants were females (n =32, 55.2%), 37.9% were 50 years and over and 43.1% had a university degree. Overall score was 78.41% for knowledge, 92.8% for perceptions and 48.3% for practices. Despite the availability and gratuity only half of the Participants reported to have carried out their COVID-19 test ever since the pandemic started. Out of 40 (69.0) participants who have heard stories concerning the vaccines, 39.7% suggested that they COVID-19 vaccines can protect us from the disease. Also, out of the 58 participants who received their first dose of the anti-COVID-19 vaccine, 36 of them came for their second shot amounting for a 62% complete vaccination rate. Headache, pain and heaviness of the injected arm were the most frequently reported side effects following participants first shot. The mobile based strategy could be the best solution to increase vaccine uptake in Cameroon.

## INTRODUCTION

The severe acute respiratory syndrome coronavirus 2 (SARS-CoV-2) responsible for the Coronavirus disease (COVID-19) is a deadly virus which continues to affect many countries in the world [1]. In Cameroon, between February 25 and March 3, 2021, the occurrence of a new wave of contaminations coupled with the re-emergence of Covid-19 cases were registered. These happenings occurred when the scientific community had a new response to this pandemic, which is the vaccination that is being introduced in many countries.

The World Health Organization (WHO), in collaboration with other partners and Governments have supported global efforts which have resulted in the rapid development of effective vaccines against Covid-19. The COVAX facility was thus established in order to guarantee equitable access to these vaccines to all countries of the world. At the regional level, the African Union initiative was also created with the ambition to cover at least 60% of the African population by 2022 thanks to a mechanism of pooling of costs and requests for vaccines between States members. At the national level, Cameroon has taken advantage of bilateral cooperation with other state partners and international institutions for better access to funding and vaccine doses.

Cameroon received her first batch of the COVID-19 vaccines: 200,000 doses Sinopharm and 391,200 doses of AstraZeneca on April 11 and 17, 2021 respectively. Subsequently, Cameroon received doses of the additional vaccines from the COVAX facility and the African Union Initiative which now allows it to meet the current demand for vaccines by relying on subsequent increasing allocations.

In the initial phase of vaccination, priority target groups consisting of health workers, people living with co-morbidities and people aged 50 and over were exclusively targeted. Subsequently, the Government decided to expand the offer of vaccination to people aged 18 and over, without changing the above-mentioned order of priority. To this end, 244 vaccination centers were selected to offer daily vaccination services in all 190 health districts and, two rounds of the vaccination intensification campaign with the deployment of mobile teams in all regions were organized.

As of September 10, 2021, 438,000 doses have been administered representing 40% of the allocation of vaccines received for the period, 358,000 people have received at least one dose of vaccine, about 2.8% of the population aged 18 and over (target population) and 104,200 people have completed their vaccination schedule, exactly 0.7% of the target population.

The observation of the implementation of the vaccine strategy to date confirms the management of vaccine hesitation as the major challenge to be taken up. This vaccine hesitation, largely fueled by the proliferation of rumors and disinformation messages conveyed through social networks, is now the main cause of the slowness of vaccination, including among health workers.

Moreover, the lessons learned from the introduction phase of vaccination show that the most effective service offer approach in the Cameroonian context remains the periodic organization of vaccination intensification campaigns to strengthen the daily supply in vaccination centers that should be extended to every 1939 Health Areas. Vaccine administration strategies have been diversified with the switch to routine vaccination in all health areas, periodic intensification through campaigns with deployment of mobile teams, and the establishment of large health centers. vaccination with logistical means adapted for mobile sessions in urban areas.

It is in this light that the Biyem assi mosque has requested a mobile team from the Ministry of Public Health. Our study aimed at determining the KAP and the epidemiological profile of Muslim faithful receiving the anti-Covid-19 vaccine at Mosquée Biyem-Assi, Yaoundé-Cameroon and evaluate the association between the demographics and KAP of Covid-19.

## METHODOLOGY

### Study setting

This cross-sectional study was carried out from June to July 2021, in the Biyem-Assi health district precisely at the Central Mosque premises. Biyem-Assi is the headquarters of the Yaounde VI Sub-division in the Mfoundi Division, Centre Region of Cameroon. It is located at latitudes 11°29’12” East and longitude 3°50’25” North. Biyem-Assi constitutes one of the most populated sub-divisions of the Mfoundi Division with about 300,000 inhabitants. It is a very vast zone that englobes several neighbourhoods like Rond-point Express, Accacias, Carrefour Biyem-Assi, Maison Blanche, Montee des Soeurs, Montee Jouvence, Superette, Tam-Tam and TKC. Yaounde VI has an equatorial climate with four seasons: two rainy seasons (a long one from March to June and short one which lasts from September to October) and two dry seasons (the long dry season runs from November to March and a short one which lasts from July to August).

### Study design

Data on COVID-19 was collected using a well-structured questionnaire established following items based on previous literature and administered face to face to all consenting participants. It covered four sections (demography, knowledge, perceptions/attitudes and practices). The questionnaire had 41 questions: 18 on knowledge, 3 on perception, 7 on practices, 11 on demographics, and 2 others (source of information and feedback information during the second dose vaccination). Questions were answered on a Yes/No/I don’t know basis. Also, Always/Occasional/Never were answer items provided for questions on practices and some open-ended questions were equally asked. Also, health parameters (temperature, BMI, blood pressure) and rumours about the vaccines were equally registered for each participant. The relations between the demographics and KAP were assessed.

### Study population and eligibility criteria

Adults from all works of life aged 18 and above who come for the COVID-19 vaccination and who agreed to participate in the study, completed the survey form.

### Study size

In this study, the sample size was calculated following the formula proposed by Lorentz, stated as: **N** = **[z^2^ × P (1-P)]/e^2^**

Where; **N**: sample size, **z**: 1.96 (value of the 95% confidence interval), **p**: standard deviation (prevalence) taken at 50% and **e**: margin of error taken at 5%.

Therefore, N = [1.96^2^×0.5 (1-0.5)] /0.05^2^ = 384.16. However, our population size was 58.

### Statistical methods

Data analyses of this study were performed using Microsoft Excel 2019 and Statistical Package for the Social Sciences (SPSS), version 25. Descriptive analysis was used to study demographics such as age, gender, education, profession as well as socio-economic status. The mean, standard deviation and range were calculated for quantitative parameters. Also, associations between demographics and KAP were studied. The statistical significance level was set at p < 0.05.

### Ethical Consideration

The study was approved by the National Ethics Committee and was conducted following the ethical principles of the World Medical Association Declaration of Helsinki. Patients’ personal identities and other private information were anonymized; therefore, the need for informed consent was waived in accordance with local legislation and national guidelines. A go ahead from the Ministry of health through a service note and full approval of this study was also granted by the religious leader (IMAM) of the Biyem-Assi Muslim community. Finally, every member signed a consent form before they were included in the study.

## RESULTS

### Socio-demographic characteristics

A total of 58 participants took part in the first survey and equally received their first dose of the anti-COVID-19 vaccine of their choice. The mean age of participants was 46 ± 15 years with 37.9% aged 50 years and over. Majority of the study participants were females (n =32 ; 55.2%), and 43.1% had a university degree. Majority of respondents live in houses with more than 6 inhabitants (Table I).

**Table I:** Demographic characteristics of participants

| **Characteristics** |  | **Frequency (%)**<br>**(n=58)** |
| --- | --- | --- |
| **Gender** | Male | 26 (44.8) |
|  | Female | 32 (55.2) |
| **Age-group (years)** | 18-35 | 15 (25.9) |
|  | 36-50 | 21 (36.2) |
|  | >50 | 22 (37.9) |
| **Marital status** | Single | 1 (3.6) |
|  | Married | 26 (99.2) |
|  | divorced | 1 (3.6) |
| **Education** | Have never been to school | 10 (17,2) |
|  | Primary | 8 (13.8) |
|  | Secondary | 15 (25.9) |
|  | University | 25 (43.1) |
| **Profession** | Healthcare worker | 3 (5.2) |
|  | Public service worker | 17 (29.3) |
|  | Private sector worker | 4 (6.9) |
|  | Teacher/lecturer | 2 (3.4) |
|  | Retired | 6 (10.3) |
|  | Student | 5 (8.6) |
|  | Others | 21 (36.2) |
| **Weight status** | Underweight | 2 (4.3) |
|  | Normal weight | 17 (36.2) |
|  | Overweight | 17 (36.2) |
|  | Obese | 11 (23.4) |
| **Standard of living** | Poor | 28 (48.3) |
|  | Average | 19 (32.8) |
|  | Rich | 11 (19.0) |
| **Number of people in households** | 1-3 | 9 (15.5) |
|  | 4-6 | 11 (19.0) |
|  | >6 | 38 (65.5) |

*Others include:* religious leaders, taxi drivers, unemployed, housewives

### Source of information regarding the COVID-19 vaccines and participants health status

Concerning the source of information regarding COVID-19 vaccines, 89.7% of the study population answered yes to have heard about the vaccines on television followed by social media platforms accounting for 75.9% yes rate, followed by the radio (72.4%), religious leaders (51.7%) and from family members and friends (46.6%) (Figure 1). Table 2 gives a clear outline of our participants’ health status. We could consider our study population to be generally of good health judging from the 48.3% who only go for medical checkups when need arises. Twenty-two percent of the participants had chronic diseases, with hypertension and asthma accounting for 33.3 % each.

**Table II:** Description of participants health status

| Characteristics | Frequency | Percentage |
| --- | --- | --- |
| **How often do you go for a health check-up?** |  |  |
| Once in 3 months | 8 | 13.8 |
| Once in 6 months | 7 | 12.1 |
| Once a year | 9 | 15.5 |
| Only when needed | 28 | 48.3 |
| Never go for one | 6 | 10.3 |
| **Do you have any chronic diseases** |  |  |
| Yes | 13 | 22.4 |
| No | 45 | 77.6 |
| **Type** |  |  |
| Asthma | 4 | 33,3 |
| headache | 1 | 8,3 |
| Diabetes | 1 | 8,3 |
| Glaucoma | 1 | 8,3 |
| Hypertension | 4 | 33,3 |
| Lower back pain | 1 | 8,3 |

**Figure 1:**
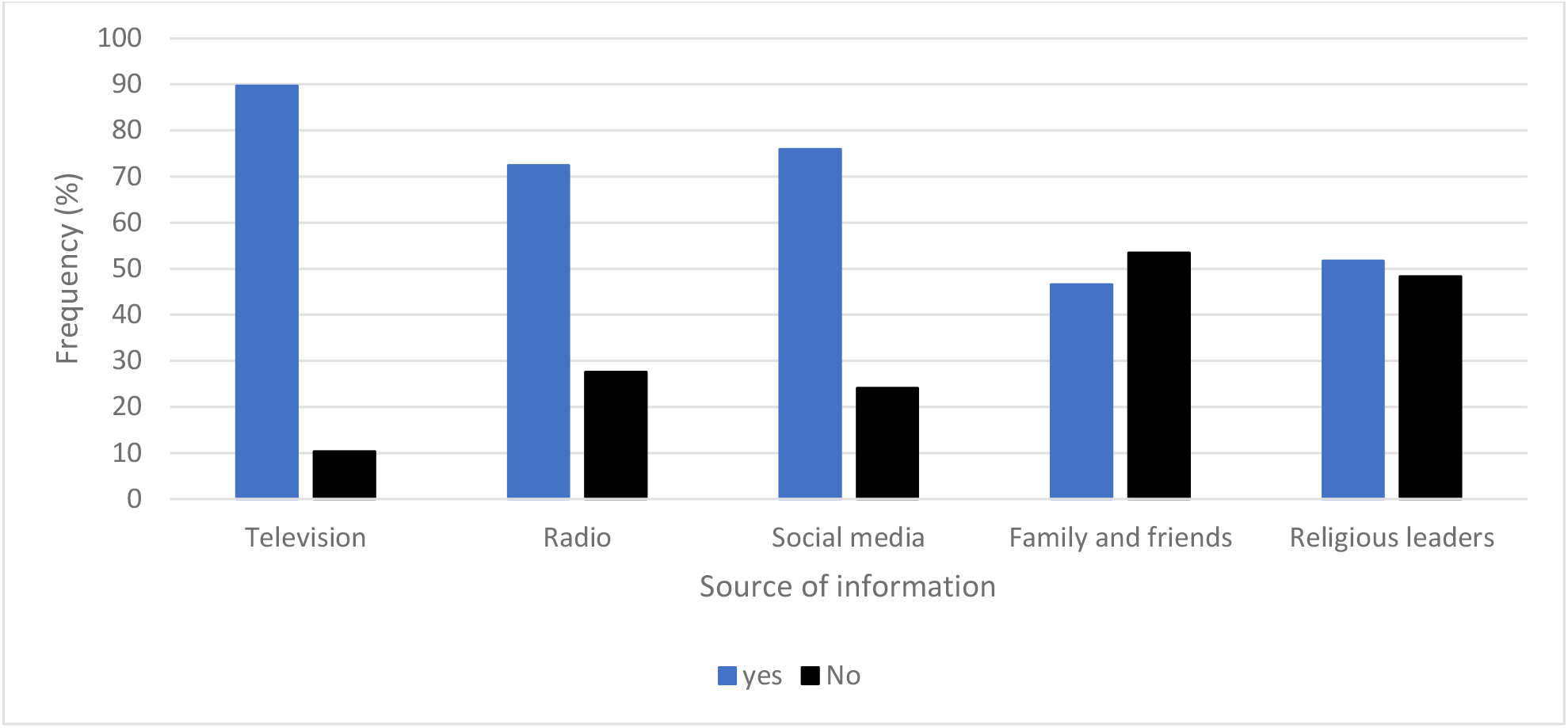
Source of information on COVID-19 vaccines

### Knowledge of participants related to COVID 19

Almost all (96.6%) of the study participants new about the coronavirus and 91.4% could correctly define it. With respect to the mode of transmission, 74.1% reported that the disease could be contracted through respiratory droplets of an infected person, 81% when in close contact with an infected person and 65.5% said through contact with infected surfaces or objects (Table 3). Overall, our results show that 78.41 (13.33/17*100) had high knowledge score of the disease (mean COVID-19 knowledge score:13.33 **±** 2.2, range: 10-17) (Table IV).

**Table III:** Knowledge, attitudes, and practices of study participants

|  | **Questionnaire items**<br>**About Covid-19** | **Responses (N = 58)** |  |  |
| --- | --- | --- | --- | --- |
|  |  | **Yes**<br>**(n, %)** | **No**<br>**(n, %)** | **I don't**<br>**know**<br>**(n, %)** |
| **Knowledge** | Have you heard about COVID-19 | 56 (96.6) | 2 (3.4) | - |
|  | COVID-19 is a contagious disease caused by the coronavirus (True) | 53 (91.4) | 5 (8.6) | - |
|  | Covid19 is a disease caused by bacteria (False) | 9 (15.5) | 49 (84.5) | - |
|  | Covid19 is a fatal disease in the absence of treatment (True) | 9 (15.5) | 49 (84.5) | - |
|  | Can everyone get infected with COVID-19? (True) | 47 (81.0) | 11 (19.0) | - |
|  | Can COVID-19 be prevented? (True) | 58 (100.0) | - | - |
|  | **Mode of transmission** |  |  |  |
|  | COVID-19 can be contracted through contact with or consumption of wild animals (False) | 49 (84.5) | 9 (15.5) | - |
|  | COVID-19 can be contracted through respiratory droplets of an infected person (True) | 43 (74.1) | 15 (25.9) | - |
|  | COVID-19 can be contracted when in close contact with an infected person (True) | 47 (81.0) | 11 (19.0) | - |
|  | COVID-19 can be contracted through contact with infected surfaces or objects (True) | 38 (65.5) | 20 (34.5) | - |
|  | **Symptoms of COVID-19** |  |  |  |
|  | Fever | 47 (81.0) | 11 (19.0) | - |
|  | Cough | 57 (98.3) | 1 (1.7) | - |
|  | Headaches and general fatigue | 48 (82.8) | 10 (17.2) | - |
|  | Difficulty in breathing | 48 (82.8) | 10 (17.2) | - |
|  | Have a flu | 30 (51.7) | 28 (48.3) | - |
| **Perceptions** | **About anti-COVID-19 vaccines** |  |  |  |
|  | Have you heard about anti-COVID-19 vaccines? | 55 (94.8) | 3 (5.2) | - |
|  | Sinopharm is an anti-COVID-19 vaccine available in the country? | 44 (75.9) | 10 (17.2) | 4 (6.9) |
|  | AstraZeneca is an anti-COVID-19 vaccine available in the country? | 46 (79.3) | 8 (13.8) | 4 (6.9) |
|  | Do you think the Covid-19 test kits are reliable? | 53 (91.4) | 3 (5.2) | 2 (3.4) |
|  | In your opinion, is it important to get tested for Covid-19? | 56 (96.6) | 2 (3.4) | - |
|  | Do you think the vaccines are effective? | 50 (86.2) | 8 (13.8) | - |
|  | Have you taken a Covid-19 test before? | 32 (55.2) | 26 (44.8) | - |
| **Practices** | **Respect of barrier measures** | **Always** | **Occasional** | **Never** |
|  | Wearing of face mask | 5 (8.6) | 23 (39.6) | 30 (51.7) |
|  | Handwashing with soap and clean water | 15 (25.9) | 13 (22.4) | 30 (51.7) |
|  | Use of hand sanitizers | 1 (1.7) | 21 (36.2) | 36 (62.1) |
|  | Respect of social distancing | - | 23 (39.7) | 35 (60.3) |
|  | Coughing into a flex elbow | 9 (15.5) | 15 (25.8) | 34 (58.6) |
|  | Taking home made concoctions | - | 11 (19.0) | 47 (81.0) |

**Table IV:** Associations between some background characters (sex, age, education) of participants and knowledge score of COVID-19/COVID-19 vaccines

| Variables | | Number of participants (%) | Knowledge score (Mean ± standard deviation) | p-value |
| --- | --- | --- | --- | --- |
| **Overall** |  | 58 (100) | 13.33 (2.2) | - |
| **Sex** | F | 26 (44.8) | 13,1 (2.1) | 0.446 |
|  | M | 32 (55.2) | 13.5 (2.3) |  |
| **Age** | 18-35 | 15 (25.9) | 13.4 (2.4) | 0.582 |
|  | 36-50 | 21 (36.2) | 13.7 (1.8) |  |
|  | >50 | 22 (37.9) | 13.0 (2.6) |  |
| **Education** | Have never been to school | 10 (17,2) | 13.0 (2.7) | 0.137 |
|  | Primary | 8 (13.8) | 11.8 (3.3) |  |
|  | Secondary | 15 (25.9) | 13.8 (2.0) |  |
|  | University | 25 (43.1) | 13.7 (1.5) |  |

### Perceptions of participants towards COVID 19

Regarding participants’ impression about the COVID 19 test kits, 91.4% thought they were reliable, 96.6% considered getting tested as important while 86.2% esteemed the available vaccines as effective (Table 3). Overall, 92.8% of respondents had a high perception score (Table V).

**Table V:** Associations between some background characters (sex, age, education) of participants and perception score of COVID-19/COVID-19 vaccines

| Variables | | Number of participants (%) | Perception score (Mean ± standard deviation) | p-value |
| --- | --- | --- | --- | --- |
| **Overall** |  | 58 (100) | 4.64 (0.7) |  |
| **Sex** | F | 26 (44.8) | 4.6 (0.6) | 0.550 |
|  | M | 32 (55.2) | 4.7 (0.7) |  |
| **Age** | 18-35 | 15 (25.9) | 4.7 (0.6) | 0.980 |
|  | 36-50 | 21 (36.2) | 4.6 (0.7) |  |
|  | >50 | 22 (37.9) | 4.6 (0.7) |  |
| **Education** | Have never been to school | 10 (17.2) | 4.4 (0.7) | 0.086 |
|  | Primary | 8 (13.8) | 4.4 (1.2) |  |
|  | Secondary | 15 (25.9) | 5.0 (0.0) |  |
|  | University | 25 (43.1) | 4.6 (0.6) |  |

### Practice of participants towards COVID 19

Participants’ practices toward COVID-19 are shown in Table 3. 32 (55.2%) participants reported to have carried out their COVID-19 test ever since the pandemic started. Results show that only 8.6% of the population always wore their face masks while 15 (25.9%) of them had the habit of always washing their hands with soap and clean water. Generally, the practice score was 48.3% (Table VI). Practice score was however significant for women (p=0.018).

**Table VI:**
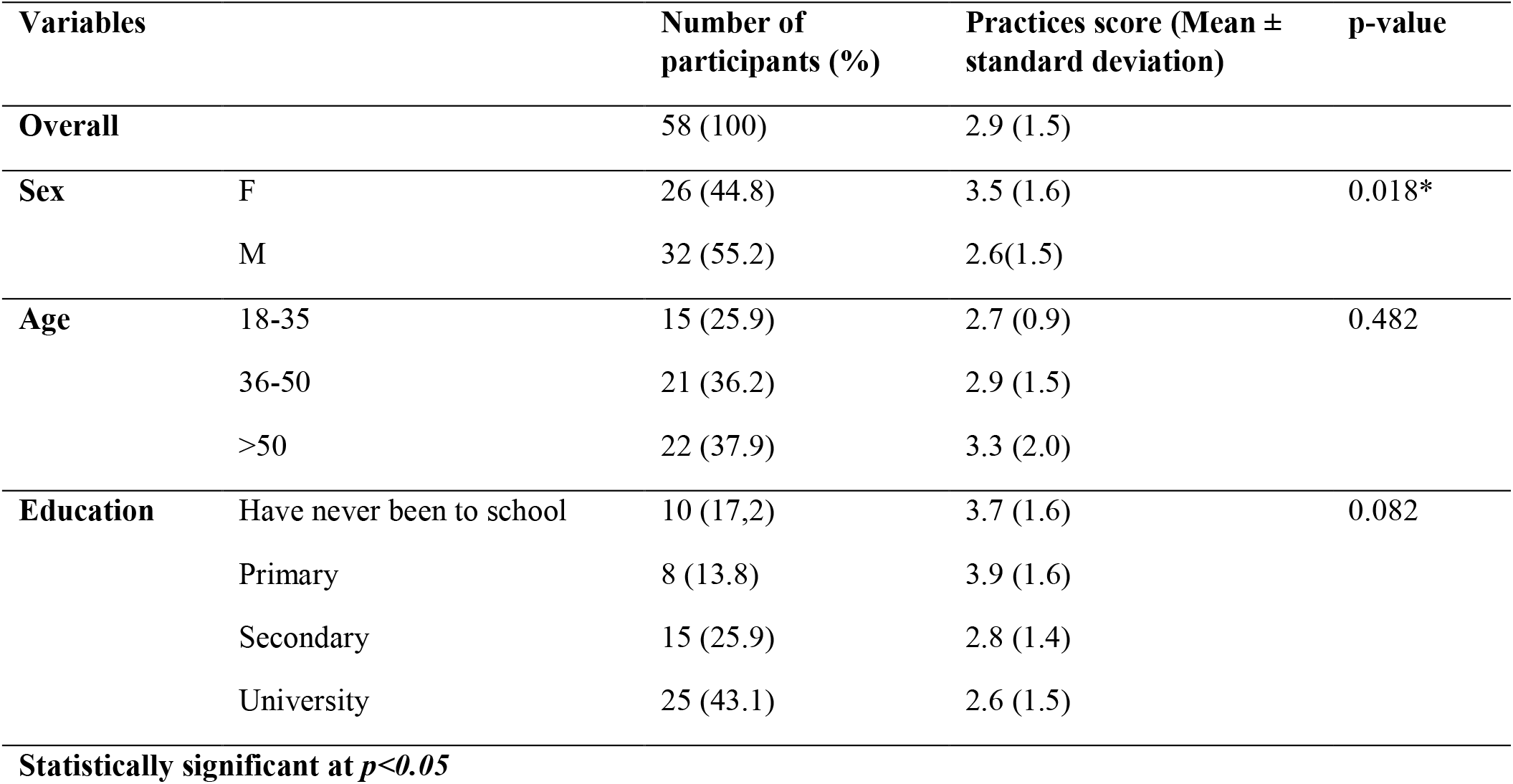
Associations between some background characters (sex, age, education) of participants and practice score of COVID-19/COVID-19 vaccines

| Variables |  | Number of participants (%) | Practices score (Mean ± standard deviation) | p-value |
| --- | --- | --- | --- | --- |
| **Overall** |  | 58 (100) | 2.9 (1.5) |  |
| **Sex** | F | 26 (44.8) | 3.5 (1.6) | 0.018* |
|  | M | 32 (55.2) | 2.6(1.5) |  |
| **Age** | 18-35 | 15 (25.9) | 2.7 (0.9) | 0.482 |
|  | 36-50 | 21 (36.2) | 2.9 (1.5) |  |
|  | >50 | 22 (37.9) | 3.3 (2.0) |  |
| **Education** | Have never been to school | 10 (17,2) | 3.7 (1.6) | 0.082 |
|  | Primary | 8 (13.8) | 3.9 (1.6) |  |
|  | Secondary | 15 (25.9) | 2.8 (1.4) |  |
|  | University | 25 (43.1) | 2.6 (1.5) |  |

Statistically significant at $p<0.05$

### Rumors/stories about the COVID-19 vaccines

out of 40 (69.0) participants who have heard stories concerning the vaccines, 39.7% suggested that they COVID-19 vaccines can protect us from the disease while some 22.4 said the vaccines have too many side effects like blood clotting, 17.2% suggested that the vaccines are not effective, 15.5% vaccines are to control humanity, 3.4% can cause sterility and 1.7% said it was a means to kill the African population (Table VII).

**Table VII:** Rumors/stories about the COVID-19 vaccines

| Characteristics | Yes (%) | No (%) |
| --- | --- | --- |
| Have you heard any rumors about the available COVID-19 vaccines | 40 (69.0) | 18 (31.0) |
| If yes, what did you hear? |  |  |
| The vaccines are a means to infect Africans | 1 (1.7) | - |
| The vaccines are not effective | 10 (17.2) | - |
| The vaccines have been made to control humanity | 9 (15.5) | - |
| The vaccines have too many side effects like blood clotting | 13 (22.4) |  |
| The vaccines can cause sterility | 2 (3.4) |  |
| The vaccines can protect us against COVID-19 | 23 (39.7) | - |

### Feedback from participants during the second dose

Out of the 58 participants who received their first dose of the anti-COVID-19 vaccine, 36 of them came for their second shot amounting for a 62% complete vaccination rate. Headache, pain and heaviness of the injected arm were the most frequently reported side effects following participants first shot. Overall, participants were very appreciative of the vaccination setting (Table VIII)

**Table VIII:** Feedback from participants during the second dose

| Questionnaire items About Covid-19 vaccine | Responses (N = 61) |  |
| --- | --- | --- |
|  | Yes | No |
|  | (n, %) | (n, %) |
| Have you received the COVID-19 vaccine? | 36 (59) | 25 (41) |
| Did you experience any side effects after your COVID-19 vaccine? | 11 (18) | 50 (82) |
| **What were the side effects?** |  |  |
| Headache | 5 (8.2) | 56 (91.8) |
| Dizziness | 1 (1.6) | 60 (98.4) |
| Pain at the site of injection | 6 (9.8) | 55 (90.2) |
| Heaviness of the injected arm | 5 (8.2) | 56 (91.8) |
| Other | 2 (3.3) | 59 (96.7) |
| Did you experience any difficulties when attending your COVID-19 vaccine appointment? | 1 (1.6) | 60 (98.4) |
| **\*Others (fatigue)** |  |  |

## DISCUSSION

Over the years, immunization has played a fundamental role in reducing the burden of infectious diseases. Consequently in 2001, some 33,000 deaths and 14 million diseases were prevented [2]. In order to halt the ongoing pandemic, several COVID-19 vaccines have been approved and are currently being administered worldwide. In Cameroon, as of October 18, 2021, exactly 189 days (half a year) after the start of the vaccination, 490,604 doses have been administered or approximately 2,600 doses on daily basis. The most used vaccine being AstraZeneca with nearly 307 674 doses administered. Within a target population of 13 944 491, 2.9% received at least one dose of vaccine and 1.1% were fully immunized (MINSANTE np). Despite the numerous immunization centers in Cameroon, COVID-19 roll-out still poses a problem. This has raised doubts about the general population’s knowledge, perceptions and practices towards the COVID-19 vaccine.

In this study, knowledge about COVID-19 vaccine and vaccination was high among study participants. Knowledge score did not vary significantly with gender, age and education. This finding is similar with other studies demonstrating no significant sex, age and education differences in knowledge regarding COVID-19 [3–5]. Nevertheless, our results differ from studies concerning knowledge towards COVID-19 carried out in Bangladesh which reported that males had marginally higher scores in knowledge regarding COVID-19 than females [6]. The good knowledge score registered in our study on COVID-19 vaccines and vaccinations is possibly due to high exposures to information or publicity on COVID-19 vaccinations following the numerous vaccination campaigns organized by the government. Also, the fact that the study was carried out in an urban area, could explain the increase in awareness due to regular access to electricity and internet which are necessary tools to connect to media platforms resulting in regular access to COVID-19 related information shared here [7]. Hence, the increase knowledge about COVID-19, transmission pathways, barrier measures and information on the available vaccines in the country.

Participant’s perception score was high 4.64 (0.7). This can be translated into the amount of knowledge they have regarding the COVID-19 vaccines effectiveness and safety. Also, the fact that the vaccination team was invited by the Muslim community of Mosquee Biyem-Assi clearly shows the amount of influence and trust that reigns in the religious arena.

Adherence to preventive practices such as the washing of hands was very appreciable compared to practice of other barrier measures. This could primarily be attributed to the Muslim religion which entails the washing of hands and feet before prayers coupled with the government order to put on face masks and washing of hands at every public place. The laxity in the respect of barrier measures could be attributed to the presence of available traditional concoctions esteem as being effective against the disease [3].

Amongst the chronic diseases reported by participants, hypertension and asthma were the most prevalent (33.3%) in respondents. This could be highly risky for these participants given that, some studies on coronavirus death rate tendency have been reported to be recurrent in persons with pre-existing comorbidity such as Hypertension, Chronic respiratory disease, cardiovascular disease, Diabetes and Cancer [8,9]. Respondents did not report any severe side effects after vaccinations this shows that the vaccines are safe and reliable. Out of the 58 participants who received the first dose, 36 of them came back for their second shot accounting for a complete immunization rate of 62%. This increase in immunization rate suggest that the mobile vaccination strategy could be the best solution at the moment to increase vaccine uptake in Cameroon.

## LIMITATIONS

The limitation of this study is the small sample size and limited sample representativeness which does not permit us draw a suitable conclusion regarding KAP.

## CONCLUSION

Overall, our analyses revealed that the mobile based strategy could be effective in the COVID-19 immunization coverage plan. This paper also reveals that the Muslim community of Mosque Biyem-Assi have a good knowledge of the disease but portray poor practices of barrier measures which is a potential risk given the current vaccination rate. Hence, the need to intensify health education programs and intensify laws on the respect of barrier measures.

## Data Availability

All data produced in the present study are available upon reasonable request to the authors

## Author Contributions

**Conceptualization:** Adidja Amani, Efietngab Atembeh-Noura.

**Data curation:** Efietngab Atembeh-Noura, CHEUYEM LEKEUMO Fabrice Zobel

**Formal analysis:** Adidja Amani, CHEUYEM LEKEUMO Fabrice Zobel, Efietngab Atembeh-Noura, Moussa Souaibou, Joseph KAMGNO, Bissek Anne Cécile.

**Investigation:** Adidja Amani, Efietngab Atembeh-Noura, Moussa Souaibou, Joseph KAMGNO, Bissek Anne Cécile.

**Methodology:** Adidja Amani, Efietngab Atembeh-Noura, Joseph KAMGNO, Bissek Anne Cécile.

**Project administration:** Adidja Amani, Efietngab Atembeh-Noura, Joseph KAMGNO, Bissek Anne Cécile.

**Supervision:** Adidja Amani, Efietngab Atembeh-Noura.

**Validation:** Adidja Amani.

**Visualization:** Adidja Amani.

**Writing – original draft:** Adidja Amani, Efietngab Atembeh-Noura, Jean Thierry Ebogo Belobo, Hillary Tandjeu.

**Writing – review & editing:** Adidja Amani, Efietngab Atembeh-Noura, Hillary Tandjeu, Jean Thierry Ebogo Belobo, Moussa Souaibou, Joseph KAMGNO, Bissek Anne Cécile.

### Competing interests

The authors declare no competing interest

## Acknowledgements

This study received no specific funding or grant from any agency in the public, commercial, or not-for-profit sectors.

## Declaration of interest statement

No potential conflict of interest was reported by the authors

## Notes

### Competing Interest Statement

The authors have declared no competing interest.

### Funding Statement

This study did not receive any funding

### Author Declarations

Ethics committee/IRB of the National Ethics Committee waived ethical approval for this work

